# Mortality of the COVID-19 outbreak in Sweden in relation to previous severe disease outbreaks

**DOI:** 10.1101/2020.05.22.20110320

**Authors:** Anders Ledberg

## Abstract

Influenza viruses have caused disease outbreaks in human societies for a long time. Influenza often has rapid onset and relatively short duration, both in the individual and in the population. The case fatality rate varies for different strains of the virus, as do the effects on total mortality. Outbreaks related to coronavirus infections have recently become a global concern but much less is known about the dynamics of these outbreaks and their effects on mortality.

In this work, disease outbreaks in Sweden, in the time period of 1860-2020, are characterized and compared to the currently ongoing COVID-19 outbreak. The focus is on outbreaks with a sharp increase in all-cause mortality. Outbreak onset is defined as the time point when death counts start to increase consistently for a period of at least 10 days. The duration of the outbreak is defined as the time period in which mortality rates are elevated. Excess mortality is estimated by standard methods.

In total there were 15 outbreaks detected in the time period, the first 14 were likely caused by influenza virus infections, the last by SARS-CoV-2. The mortality dynamics of the SARS-CoV-2 outbreak is shown to be similar to outbreaks due to influenza virus, and in terms of the number of excess deaths, it is the worst outbreak in Sweden since the ‘Spanish flu’ of 1918-1919.

## 1 Introduction

Influenza viruses of type A are known to have caused disease outbreaks at least since 19th century, and probably for much longer (Potter 2001). The severity of the disease caused by an influenza virus infection depends both on the properties of the virus (which strain) and on the acquired immunity and general health status of the infected individual (e.g., Cobey & Hensley 2017). The great majority recover completely from an infection, however, each year a number of persons die from consequences of influenza infections. The death toll at the level of the population depends on vaccination programs and nonpharmaceutical interventions aiming to reduce the spread of the virus (Bell et al. 2006).

Outbreaks caused by coronaviruses are thought to be a more recent phenomenon; the first reported outbreak was caused by SARS-CoV-1 in 2003 (Peiris et al. 2003), and the most recent is the still ongoing pandemic caused by SARS-CoV-2 (Huang et al. 2020). Vaccines against coronaviruses are still under development (Amanat & Krammer 2020) and measures available to control the outbreaks have so far been limited to nonpharmaceutical interventions.

It has long been recognized that influenza outbreaks often are associated with an increase in all-cause mortality that exceeds the increase directly attributed to influenza and pneumonia (e.g., Collins 1932, Reichert et al. 2004). Indeed, influenza seasons, and outbreaks, can be reliably detected from all-cause mortality data (e.g., Collins 1932, Simonsen et al. 1997). In fact, many countries and regions of the world monitor influenza by, among other things, detecting when, and by how much, the number of deaths per week exceed a preset, model based, threshold. Outbreaks such as the 1918-1920 influenza pandemic are characterized by a high attack rate and often lead to a rapidly increasing number of deaths during a short time period. Consequently, with access to daily death counts, it should be possible to detect outbreaks by looking at the local rate of change of the number of recorded deaths. Here such an approach is developed and applied to daily death counts from Sweden in the time period of 1860-2020. The excess mortality caused by the ongoing outbreak of COVID-19 is related to the 14 most severe outbreaks during the previous 160 years.

## 2 Methods

### 2.1 Data

Two sources of daily counts of deaths from all causes were used. For the years 1860-2014, data were obtained from ‘Swedish Book of Death’ issued by the The Federation of Swedish Genealogical Societies (Sveriges Släktforskarförbund 2017). This is a database compiled from a range of official sources and contains information on times and places of births and deaths for persons that have died in Sweden since 1860. The coverage is almost complete. Mortality data from 2015 until 31th of August 2020 was obtained from the website of Statistics Sweden (www.scb.se) on November 9th 2020. Data on total population size for the years 1859 to 2019 were obtained from Statistics Sweden (www.scb.se). The complete time series used in this work is available on GitHub: https://github.com/aledberg/outbreaks

### 2.2 Outbreak detection

Outbreaks were detected by analyzing the rate of change (time derivative) of daily death counts. A period of a rapid increase in death counts corresponds to a period where the derivative is consistently positive. Since the mortality data were relatively noisy, the daily deaths counts were first smoothed with a 21-point truncated Gaussian kernel and the time derivative was approximated by the first-order difference applied to the smoothed time series.

Time intervals, ten days or longer, where the rate of change exceeded a threshold value of 2.9 were selected as candidate outbreaks. The onset of the outbreak was defined as the first time point where the derivative exceeded the threshold. The offset of the outbreak was defined as the time point at which the derivative returned to zero from below. This procedure would, in principle, accurately detect the onset and offset of an outbreak described by a smooth function with a single local maximum^1^. The values of the three parameters involved in this procedure: i.e, width of the Gaussian smoothing kernel (standard deviation 1.5), the threshold value of the derivative (2.9), and the minimum number of consecutive days (10), were determined by applying the procedure to data from 1860 to 2017, i.e., not using data from the COVID-19 outbreak. Two of the outbreaks detected using this method, both occurring before 1886, were after visual inspection determined not to qualify as outbreaks (the mortality did not exceed the background level). In some cases, the offsets of the outbreaks needed to be adjusted manually since the automatic detection based on the derivative sometimes led to an overestimate the duration, and sometimes (for the 1918-1919 outbreak) underestimated the duration due to the presence of multiple local maxima.

### 2.3 Excess mortality estimation

Excess mortality caused by a disease outbreak is usually defined as the observed number of deaths minus the expected number of deaths. To estimate the expected number of deaths a variant of the regression method first described by Robert Serfling (Serfling 1963) is often used, and this approach was also adopted here. This consists in fitting a regression model to data where time periods corresponding to the outbreaks have been removed. The model is then used to predict (forecast) values for the time period corresponding to the outbreak, and excess mortality is taken as the difference between observed death counts and counts predicted from the model.

In this work, the number of deaths per day, *N*_*t*_, was assumed to follow a Poisson distribution with a time-dependent expected value obeying the following model

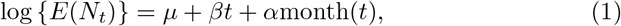

where month(*t*) is a categorical variable denoting the month corresponding to time *t*, and *µ, β*, and *α* are the parameters used to fit the model to data. Note that time, *t*, is expressed in units of days. Separate models for each outbreak were fitted to data from five years prior to the onset of the outbreak.

Excess mortality during the outbreaks was then estimated as the sum of the differences between observed and expected mortality for the days of the outbreak. Since the Swedish population has increased substantially over the time period, excess mortality was also expressed in terms of per 100,000 population. The number for the total population size was taken as the population the last of December the year before the onset of the outbreak.

Supplemental figure S1 illustrates the method used to detect outbreaks and estimate excess mortality.

### 2.4 Classification of outbreaks

To investigate if influenza virus infections might have caused the detected outbreaks, official Swedish records reporting on causes of death for the corresponding years were used. For the years prior to 1911 this information was published in the annual publication “Bidrag till Sveriges officiella statistik. A. Befolkningsstatistik (BiSOS A)”, for the years 1911-1996, causes of death were published annually in “Dödsorsaker” both issued by Statistics Sweden and available online on their website www.scb.se.

## 3 Results

Fifteen outbreaks were detected in the 160 years of mortality data analyzed (Fig 1). These outbreaks correspond to the 15 highest peaks in the data. The method used to estimate excess mortality is illustrated in Fig 2, which also shows the COVID-19 outbreak in more detail. Data characterizing the 15 outbreaks are tabulated in Table 1. In terms of excess deaths, the COVID-19 outbreak (last outbreak) is the worst since the outbreak in 1918-1919, and when standardized by the total population size, it is the worst outbreak since 1931.

**Table 1:** Outbreaks detected in the data. Excess mortality is expressed as the difference between observed and expected deaths (see Methods), and is show both in absolute numbers as well as standardized to the size of the total population.

| onset | offset | duration (days) | excess mortality | excess mortality (per 10<sup>5</sup> pop.) | common name |
| --- | --- | --- | --- | --- | --- |
| Dec 21 1889 | Feb 13 1890 | 54 | 5011 | 105.5 | 'Russian flu' |
| Dec 16 1891 | Apr 14 1892 | 120 | 7186 | 150.2 | 'Russian flu' |
| Dec 29 1893 | Feb 13 1894 | 46 | 2664 | 55.4 | 'Russian flu' |
| Dec 22 1914 | Mar 04 1915 | 72 | 5058 | 89.7 |  |
| Sep 08 1918 | Apr 25 1919 | 229 | 39391 | 679.1 | 'Spanish flu' |
| Jan 11 1922 | Mar 23 1922 | 71 | 3527 | 59.2 |  |
| Jan 21 1927 | Apr 03 1927 | 72 | 4587 | 75.5 |  |
| Jan 04 1931 | Mar 15 1931 | 70 | 4571 | 74.4 |  |
| Dec 29 1940 | Mar 01 1941 | 62 | 3145 | 49.6 |  |
| Dec 20 1950 | Feb 10 1951 | 52 | 2363 | 33.8 |  |
| Dec 15 1969 | Jan 25 1970 | 41 | 1345 | 17.0 |  |
| Nov 22 1988 | Jan 19 1989 | 58 | 2997 | 35.6 |  |
| Nov 22 1993 | Jan 24 1994 | 63 | 2877 | 33.1 |  |
| Dec 13 1995 | Jan 20 1996 | 38 | 2543 | 28.8 |  |
| Mar 19 2020 | May 31 2020 | 73 | 5216 | 50.5 | COVID-19 |

**Figure 1:**
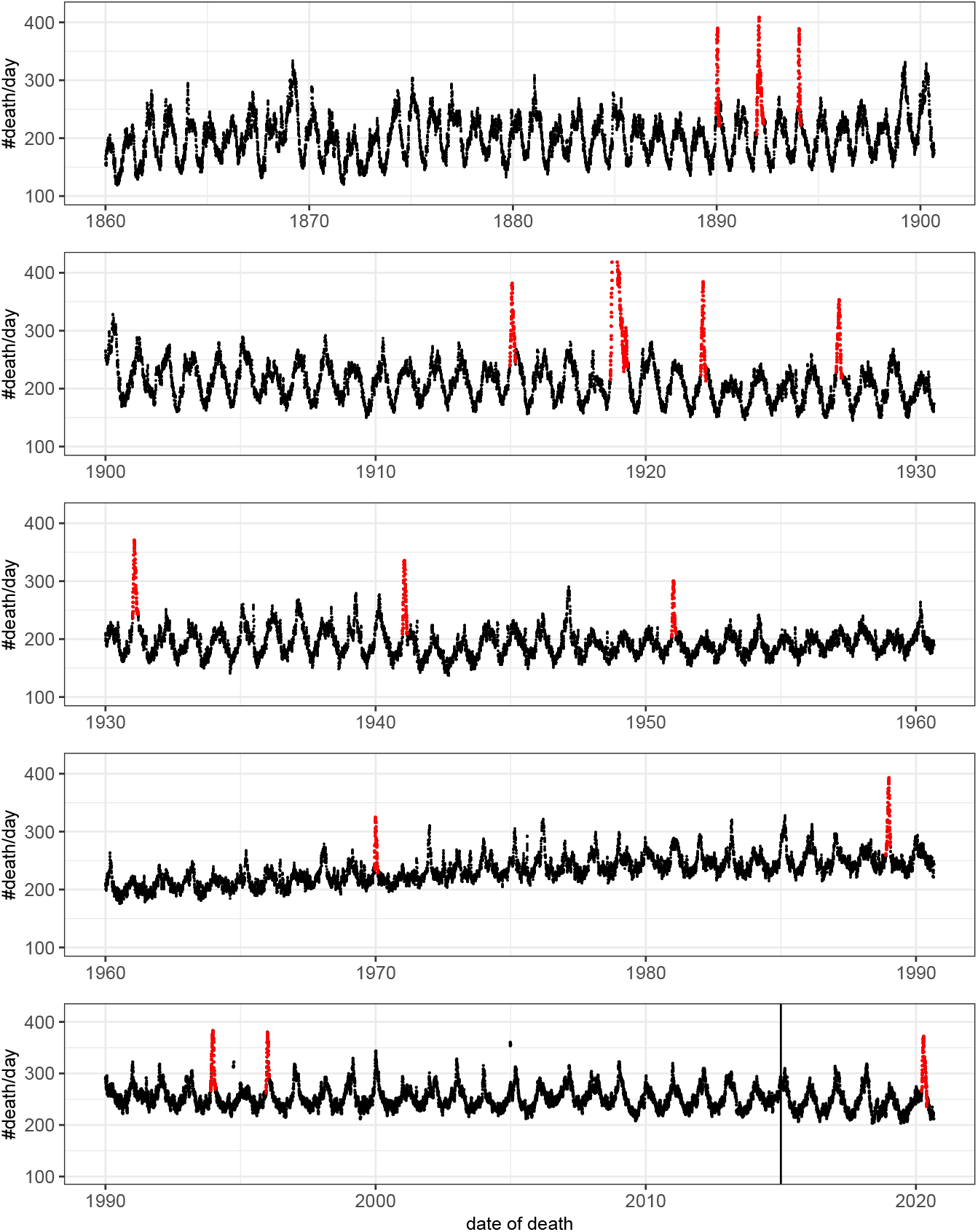
Number of deaths per day as a function of time. For visualization the original count data have been smoothed by rectangular window of length seven. That is, each value represent the mean value of the neighboring seven values. Detected outbreaks are shown in red. Note that the peak of the outbreak of 1918-1919 has been truncated. The vertical line indicates the time point at which there was a change in data source (see Methods).

**Figure 2:**
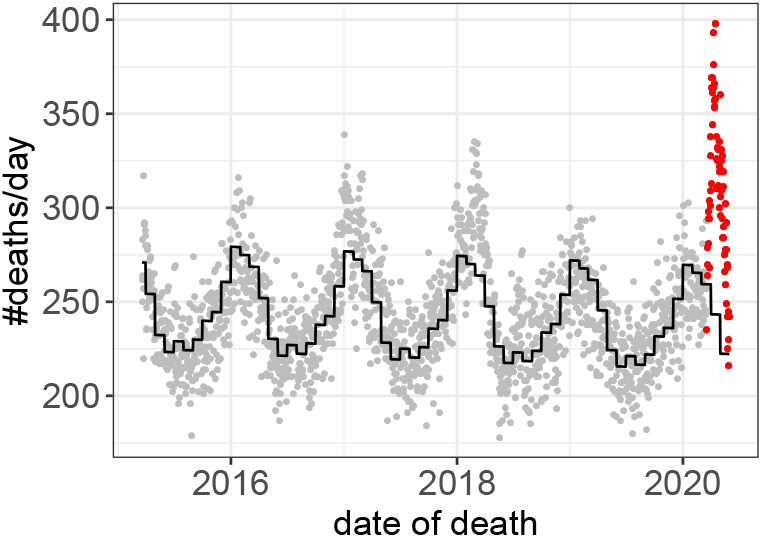
Death counts prior to, and during, the COVID-19 outbreak. Observed values prior to the outbreak in gray. Values during the outbreak in red. The predicted values based on the Poisson model (Eq 1) are shown in black.

## 4 Discussion

Using daily death counts from 1860 until present, 15 disease outbreaks, characterized by rapidly increasing all-cause mortality, were detected. Official Swedish records on causes of death (see Methods) clearly indicate that the outbreaks occurring before 1960 all coincided with influenza epidemics or pandemics. Four of these outbreaks coincided with previously characterized pandemics, see Table 1, but in most cases the influenza virus strains causing the outbreaks are not known with certainty.^2^ Likely, the outbreaks between 1960 and 2000 were also caused by influenza virus, but here the official records give less clear support. The last outbreak detected was caused by SARS-CoV-2. In terms of all-cause mortality the time-course is similar for all the 15 outbreaks; a rapid onset and a slower return to baseline. Note that the algorithm used to detect outbreaks was fine-tuned using data not including the COVID-19 outbreak, and that this outbreak was readily detected demonstrate that the dynamics is similar to outbreaks caused by influenza viruses. Most outbreaks were relatively short in duration; all except two were less than three months long. The 1918-1919 outbreak (part of the ‘Spanish flu’ pandemic) was exceptional both in excess mortality and duration, and lasted for more than half a year.

A disease outbreak might be reasonably defined as a sudden increase in the number of cases of the disease. Consequently, the outbreaks detected in this work are, of course, just a subset of all outbreaks in Sweden during the time period. Many disease outbreaks are not associated with a substantial increase in mortality rates and such outbreaks cannot be detected using mortality data. Furthermore, by looking at all-cause mortality from the entire Swedish population, local disease outbreaks, even with a marked increase in mortality, might be hard to detect. The focus, in this work, on outbreaks with rapid increases in death counts was partly a consequence of the available data. Indeed, the most conspicuous outbreaks present in the data were of this kind (Fig 1). However, outbreaks having less rapid onsets, smaller peaks, and longer durations might not be detected with the approach used here, even if their contributions to the total death count would be of comparable magnitudes. Note that the singular mortality increases caused by the sinking of MS Estonia the 28th of September 1994 and by the Indian Ocean tsunami of 26th of December 2004 were not classified as outbreaks by the method, even if the casualties, more than 500 at each occasion, led to a very sharp increase in the number of deaths around these dates. Taken together, this shows that using information local in time it is possible to reliably detect onsets of disease outbreaks from all-cause mortality data. This approach might have advantages compared to model-based approaches that defines an epidemic threshold based on data from, in some cases, several years in the past (e.g., Serfling 1963). Furthermore, when outbreaks occur outside of the classical influenza season (as was the case with COVID-19), a method not requiring a predefined time interval is beneficial.

There is no clear threshold at which an increase in deaths become an “outbreak”; changing the three parameters of the method would lead to more (or fewer) peaks being so classified. Data and code are publicly available https://github.com/aledberg/outbreaks, and the curious reader can easily try out other parameter combinations. It should be emphasized that the results obtained with respect to excess mortality are not very sensitive to the exact delimitation of the outbreak: when the observed death counts return to the expected counts, the contribution to the excess mortality is minor.

Of the eleven outbreaks that were detected in the 20th century the five first were the most severe in terms of cases per 100,000 population, and they all occurred before 1932. This decrease in severity likely has several causes, including better treatment of those infected as well as the development of influenza vaccines. It is interesting to note that the amplitude of the overall seasonality of deaths decreased under the same time period (Ledberg 2020), supporting the notion that infectious diseases are one main driver of the seasonal fluctuations in mortality (Reichert et al. 2004). The SARS-CoV-2-related outbreak in 2020 seems to be an exception from this trend of decreasing severity. In terms of absolute number of excess deaths this outbreak is the most severe since the Spanish flu in 1918-1919. Furthermore, the COVID-19 pandemic is far from over, and the final number of excess deaths will likely be much higher than the 5200 reported here.

## Data Availability

Data and R-code are available at GitHub

https://github.com/aledberg/outbreaks

## Acknowledgments

I thank Sveriges Släktforskarförbund for letting me use data from Sveriges Dödbok and Jenny Öqvist for helpful comments on previous versions of the text.

## Footnotes

1 Such a shape would result, for example, from simple compartmental models such as the SIR-model if the death counts are assumed proportional to the number of infectious people.

2 There is still a debate on what virus actually caused the 1890 pandemic. The Swedish records at the time classified this as “influenza” but this of course was based on the symptoms of the disease, and not on an identification of the causal agent.

