## Supplemental Figure 1 for "Mortality of the COVID-19 outbreak in Sweden in relation to previous severe disease outbreaks"

### Supplemental material for: Mortality of the COVID-19 outbreak in Sweden in relation to previous severe disease outbreaks

Anders Ledberg

Department of Public Health Sciences, Stockholm University
SE-106 91 Stockholm, Sweden


December 3, 2020

Figure S1: (On next page.) Illustration of the outbreak detection and excess mortality estimation. **A**: Daily number of death as reported by Statistics Sweden (black dots) and deaths smoothed with a truncated Gaussian kernel (yellow line). Standard deviation of filter kernel was 1.5 and the window was 21 points wide. **B**: Time derivative (first order difference) of smoothed death counts (black dots). Dashed line shows the threshold used for outbreak detection (2.9) and red dots show the point classified as belonging to the outbreak. Any set of 10 or more contiguous points all exceeding the threshold was taken as an indication of an outbreak. If such a set was found, the derivative was followed forward in time from it's minimum, and the outbreak was considered over when the derivative reached zero again. **C**: Black dots same as in **A**. Red dots show death counts for the days classified as belonging to the outbreak. The green curve shows the model fit based on the data prior to the outbreak, and the prediction based on the model for the period of the outbreak. **D**: Enlargement of time period close to the outbreak. Symbols have same meaning as in **C**. Excess mortality was estimated as the sum of the differences between the observed values (i.e., red points) and the predicted values (green line).

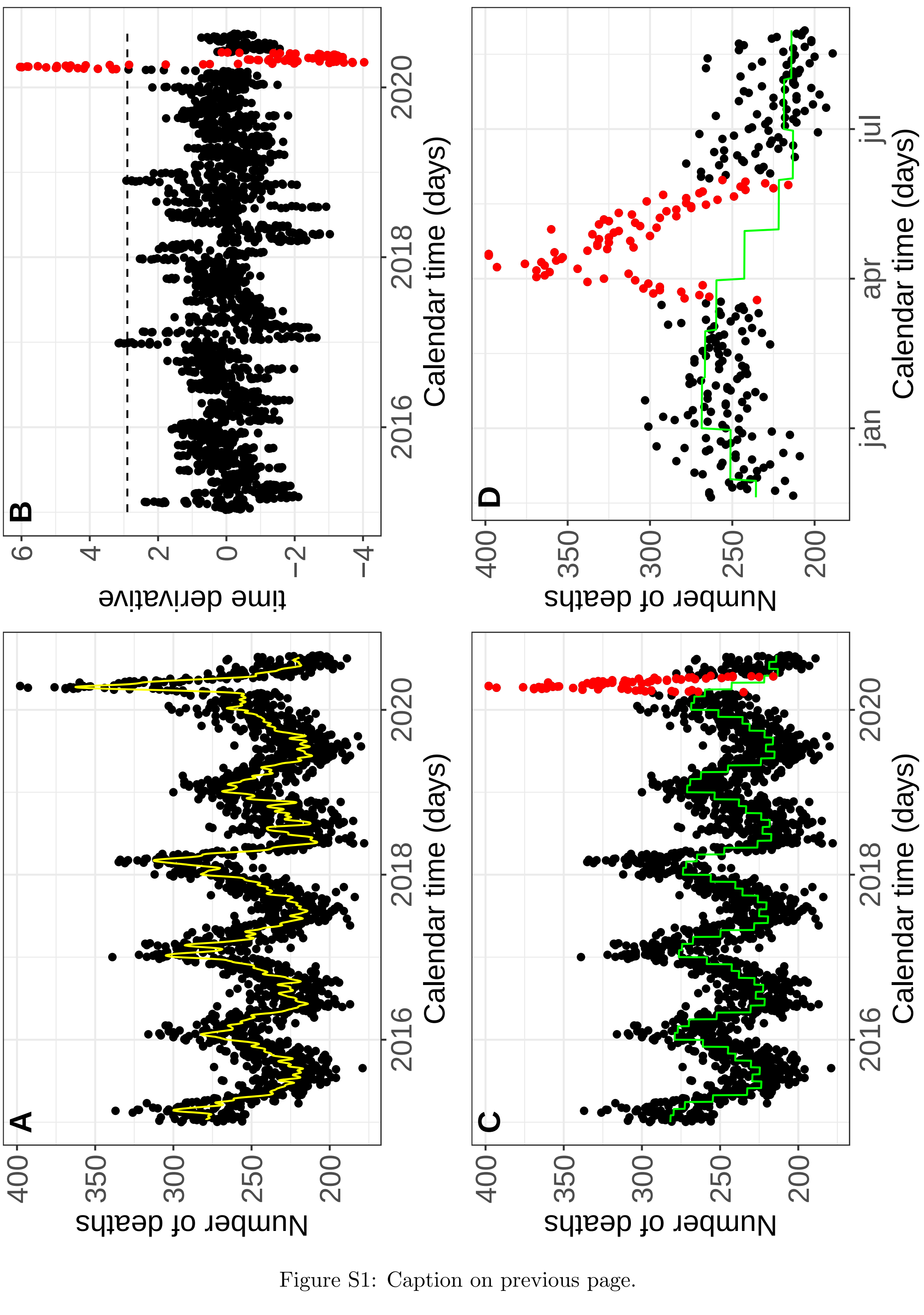


Figure S1: Caption on previous page.
